# Years Lived without Chronic Diseases after Statutory Retirement – A Register Linkage Follow-up Study in Finland 2000–2021

**DOI:** 10.64898/2026.04.12.26348889

**Authors:** Olli Pietiläinen, Aino Salonsalmi, Ossi Rahkonen, Eero Lahelma, Tea Lallukka

## Abstract

**Key points:** 1) Women get to spend more healthy years on retirement, but no clear occupational class gradient could be seen.

2) Retiring early to statutory retirement is associated with more healthy years on retirement.

3) Policies aiming to change the retirement age should consider the equitability and effects on the health of the retirees.

**Objectives:** Longer lifespans lead to longer time on retirement, despite the efforts to raise the retirement age. Therefore, it is important to study how the retirement years can be spent without diseases. This study examined socioeconomic and sociodemographic differences in healthy years spent on retirement.

**Methods:** We followed a cohort of retired Finnish municipal employees (N=4231, average follow-up 15.4 years) on national administrative registers for major chronic diseases: cancer, coronary heart disease, cerebrovascular disease, diabetes, asthma or chronic obstructive pulmonary disease, dementia, mental disorders, and alcohol-related disorders. Median healthy years on retirement and age at first occurrence of illness (ICD-10 and ATC-based) in each combination of sex, occupational class, and age of retirement were predicted using Royston-Parmar models. Prevalence rates for each diagnostic group were calculated.

**Results:** Most healthy years on retirement were spent by women having worked in semi-professional jobs who retired at age 60–62 (median predicted healthy years 11.6, 95% CI 10.4–12.7). The least healthy years on retirement were spent by men having worked in routine non-manual jobs who retired after age 62 (median predicted healthy years 6.5, 95% CI 4.4–9.5). Diabetes was slightly more common among lower occupational class women, and dementia among manual working women having retired at age 60-62.

**Discussion:** Healthy years on retirement are not enjoyed equally by women and men and those who retire early or later. Policies aiming to increase the retirement age should consider the effects of these gaps on retirees and the equitability of those effects.

## Introduction

The population is ageing in most countries, and while longer lifespans can mean more years in good health for some groups, the lengthening lifespans are globally associated with extended periods of poor health and frailty (GBD 2015 DALYs and HALE Collaborators, 2016).

Increasing lifespans mean increasing years on retirement, provided the retirement age stays the same. A study from Finland found that women are expected to spend more years on retirement than men, the difference ranging between 4.3 to 7.6 years between 1991 and 2020 (Junna et al., 2024). The same study found that expected years spent on statutory retirement are higher among men with high education and high occupational class compared to men with no qualifications or manual men respectively, but lower among women with high education compared to women with no qualifications (Junna et al., 2024). No consistent occupational class differences were observed among women. Many countries have tried to offset the fiscal and labour market effects of the ageing of the population and the diminishing young workforce by raising the retirement age. As the years spent on retirement are increasing (*Society at a Glance 2019*, 2019), it becomes crucial to ensure that these additional years are spent without diseases as long as possible. To our knowledge, no previous studies have examined healthy years on retirement.

For individuals and groups, retirement can have both positive and negative consequences. Retirement has consistently been found to have positive effects in terms of decreased use of health care, less depression and better self-reported health, while having negative effects in terms of a decline in cognition and an increase in mortality, without taking account of route of retirement (Garrouste & Perdrix, 2022). The effects of retiring early or late relative to the expected, preferred or mandated retirement age may give us indications for the effects of policies changing the retirement age on the individual. Importantly, late retirement may lead to a shorter period of healthy retirement due to the natural decline in health with age, if late retirement does not have other effects compensating for this decline. Earlier studies show little or no impact of postponed retirement on mortality, morbidity or self-reported health (Garrouste & Perdrix, 2022). There are indications that the effects of retirement may be heterogenous: the effects of retirement appear to be more commonly significant among men than women, and retirement seems to be especially beneficial for manual workers and those less educated (Garrouste & Perdrix, 2022).

Major chronic diseases such as cardiovascular diseases (Ma et al., 2023), diabetes (Heald et al., 2020), and cancers (Global Burden of Disease 2019 Cancer Collaboration, 2022) significantly reduce the number of healthy life years, but it is not known how much they reduce healthy years in retirement. Importantly, the prevalence of these diseases is not uniform across different socioeconomic groups and sexes as their occurrence is strongly socially patterned (Dalstra et al., 2005). Previous studies have consistently found education, income and wealth to be associated with healthy ageing (Wagg et al., 2021). Those in lower occupational classes have been found to have lower life expectancy, as well as lower expectancy of years lived without chronic diseases (Head et al., 2019).

Policies that encourage later retirement should consider the potential trade-offs between additional working years and the positive and negative concerns related to retirement for individuals and groups. When considering policies increasing the age of retirement, it is crucial that such policies are based on understanding whose lives they affect and how, and whether the results are equitable.

This study aims to identify factors that are associated with spending years in statutory retirement without chronic diseases. Additionally, it seeks to identify what chronic illnesses are most prevalent among those who have poor health after statutory retirement.

## Methods

### Study Population

The study population consists of employees of the City of Helsinki, Finland, first identified from the employer’s registers between 2000 and 2011 and then linked to the register of Finnish Centre for Pensions for statutory retirement using a unique personal identification number, assigned to each citizen at birth (Haukka, 2004). Employees having entered statutory retirement between 2000 and 2011 at age 60 or older were included in the study. Other routes out of employment, such as disability retirement, were not included. Those who already had an illness identified in the registers during the year preceding retirement were excluded (n=1820). After linkages, 4231 retired people were included in the study population, and were followed up on the registers for the first occurrence of illness or until the end of 2021.

Of the participants 73% were women, around one third managers and professionals and routine non-manual employees each, and around one fifth manual workers and one sixth semi-professionals (table 1). Around 37% retired at age 60-62. The average follow-up time was 15.4 years.

**Table 1:** Characteristics of the study population at the time of retirement.

|  |  |
| --- | --- |
| Age at retirement, mean (sd) | 62.8 (1.7) |
| Sex, n (%) |  |
| Women | 3091 (73) |
| Men | 1140 (27) |
| Occupational class before retirement, n (%) |  |
| Managers and professionals | 1545 (37) |
| Semi-professionals | 701 (17) |
| Routine non-manual workers | 1142 (27) |
| Manual workers | 843 (20) |
| Retirement age |  |
| 60-62 | 1553 (37) |
| 63 or over | 2678 (63) |
| Total N | 4231 |

### Measures of illness

The study population was followed on registers for the onset of illness until the end of 2021. The study population was linked to national registers on hospital discharge by the Finnish Institute for Health and Welfare (THL), prescription medicines by the Finnish Social Insurance Institution (Kela), diagnosed cancers by the Finnish Cancer Registry, and causes of death by Statistics Finland.

Eight major diagnostic groups, drawing largely on previous studies (Nyberg et al., 2020, 2022), were identified by the registers: cancer (any cancer in cancer registry, ICD-10 codes C00–C97 and D00–D09 in the hospitalization and causes of death registers), coronary heart disease (ICD-10 codes I20–I25 in the hospitalization and causes of death registers), cerebrovascular diseases (ICD-10 codes I60–I69, G45.0, G45.1, G45.2, G45.3, G45.8, G45.9 in the hospitalization and causes of death registers), diabetes (ICD-10 codes E10–E14 in the hospitalization and death registers, and ATC-codes A10A, A10B, and A10X in the medication register), asthma or chronic obstructive pulmonary disease (COPD) (ICD-10 codes J44, J45 and J46 in the hospitalization and death registers, and ATC-code R03 in the medication register), dementia (ICD-10 codes F00–F03 in the hospitalization and death registers, and ATC code N06D in the medication register), mental disorders (ICD-10 code F in the hospitalization and death registers, and ATC-codes N05 and N06 except N05C and N06D in the medication register), and alcohol-related disorders (ICD-10 codes F10, G31.2, G40.51, G62.1, G72.1, I42.6, K29.2, K70.0, K70.1, K70.3, K70.4, K86.00, K86.08, K85.2, G70.2, E24.2, T51.0 and X45 in the medication register). The first event identified in any of the registers and diagnostic groups was considered to end the healthy years on retirement for that individual.

### Factors contributing to the healthy years on retirement

Sex, occupational class, and age at retirement were used as potential factors contributing to the healthy years on retirement. Sex was based on the employer’s register, taken from the last employment contract before retirement. Occupational class was obtained from the employer’s register using the latest information before retirement, and age at retirement was calculated based on the date of retirement from the Finnish Centre for Pensions register and date of birth from the personal identification number. Sex was categorized to women and men. Occupational class was based on the occupational title sourced from the employer’s register, and was categorized to four hierarchical occupational classes: 1) managers and professionals, requiring university-level qualifications or classified as managerial positions, involving mainly autonomous managerial and supervisory tasks, and including titles such as doctors or teachers; 2) semi-professionals, requiring college-level qualifications or involving both supervisory and routine tasks with less autonomy, including titles such as nurses, foremen and technicians; 3) routine non-manual employees, requiring vocational training or no specific qualifications, involving non-supervisory clerical and other non-manual tasks, including titles such as child minders and health care assistants, and; 4) manual workers, requiring vocational training or no specific qualifications, and including those working in for example transportation or cleaning. Age at retirement was dichotomized to age 60-62 and 63 or more, based on the beginning of the eligible age of old-age retirement of 63 at 2005.

### Statistical methods

The Royston-Parmar flexible parametric survival models were used to model the baseline cumulative hazard and predict healthy years on retirement and the age of the first occurrence of illness, using duration of the time from retirement to illness as the time scale in the first case, and age as the time scale in the latter (Royston & Parmar, 2002). Unlike the Cox proportional hazards model, the Royston-Parmar model directly estimates the baseline hazard function and therefore allows prediction of the absolute measure of effect (Ng et al., 2018). Multiple models using a restricted cubic spline on the cumulative hazard scale were tested with different numbers of knots in the splines, and the best model was chosen based on the Akaike Information Criterion and visual inspection of the correspondence of the model prediction and the Kaplan-Meier curve, resulting in the choice of a model with three knots for the analysis on healthy years on retirement, and four knots for the analysis on the age of the first occurrence of illness, except in one subgroup where the four knot model could not be estimated and a two knot model was used. The models were fitted separately for all combinations of occupational class, sex, and age at retirement. Based on the estimated baseline hazard functions, median survival times and median age at the occurrence of illness along with 95 % confidence intervals (CI) were calculated from the models. Prevalence rates of the diagnostic groups were calculated in the combinations of the contributing factors. The survival modelling was performed with the flexsurv package (Jackson et al., 2024) and the analyses were conducted in the R statistical software version 4.3.1 (R Core Team, 2023).

## Results

Median healthy years on retirement and median age of first occurrence of illness calculated from survival models estimated for each combination of sex, occupational class, and age at retirement are presented in Figure 1. Women having worked in routine non-manual occupations and who retired at age 60–62 enjoyed the largest number of healthy years on retirement, 11.6 years (95% CI 10.4 to 12.7). This was followed by women having worked in semi-professional occupations at age 63 or over. Among men the largest number of healthy years on retirement were spent by semi-professionals who retired at age 60–62, with 9.6 healthy years on retirement (95% CI 5.9 to 13.9), followed by managers and professionals who retired at age 60–62.

**Figure 1:**
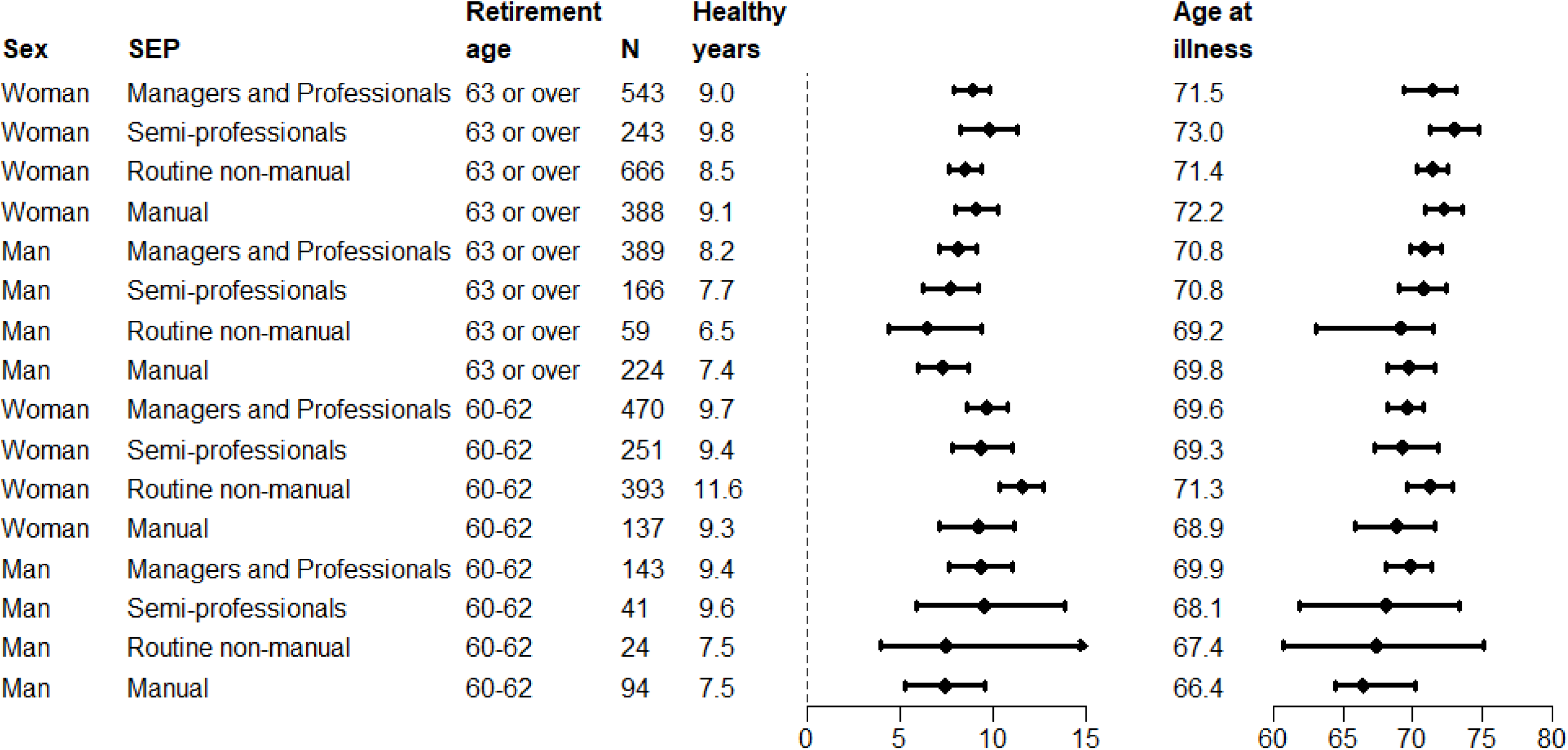
Association of sex, occupational class, and age at retirement with predicted median years lived on statutory retirement without major chronic diseases, and predicted median age at first occurrence of illness. Numeric confidence intervals can be seen in Appendix 1.

The lowest number of healthy years on retirement were spent by men retiring at age 63 or more. The lowest number of healthy years were enjoyed by men having worked in routine non-manual occupations, at 6.5 healthy years on retirement (95 % CI 4.4 to 9.5).

Overall, women consistently had more healthy years on retirement than men, and those who retired earlier had more healthy years than those who retired later. For example, the difference in healthy years on retirement between routine non-manual women who retired at age 60-62 and those who retired later was 3.1 years. No clear occupational class gradient could be seen.

Looking at the age at the first occurrence of illness, women generally got ill later in their retired life, and those who retired later also got ill later. No clear occupational class gradient was seen for the age at the first occurrence of illness.

Prevalence rates for the diagnostic groups of illness by occupational class and age at retirement can be seen in Figure 2 for women. The most common diagnostic groups among women were asthma or COPD, cancers, and mental disorders, which were observed quite equally by around a third of the women regardless of retirement age. The prevalence rates of the diagnostic groups were largely similar across occupational classes, but diabetes was somewhat more common among women from lower occupational classes regardless of retirement age and dementia among manual working women having retired early.

**Figure 2:**
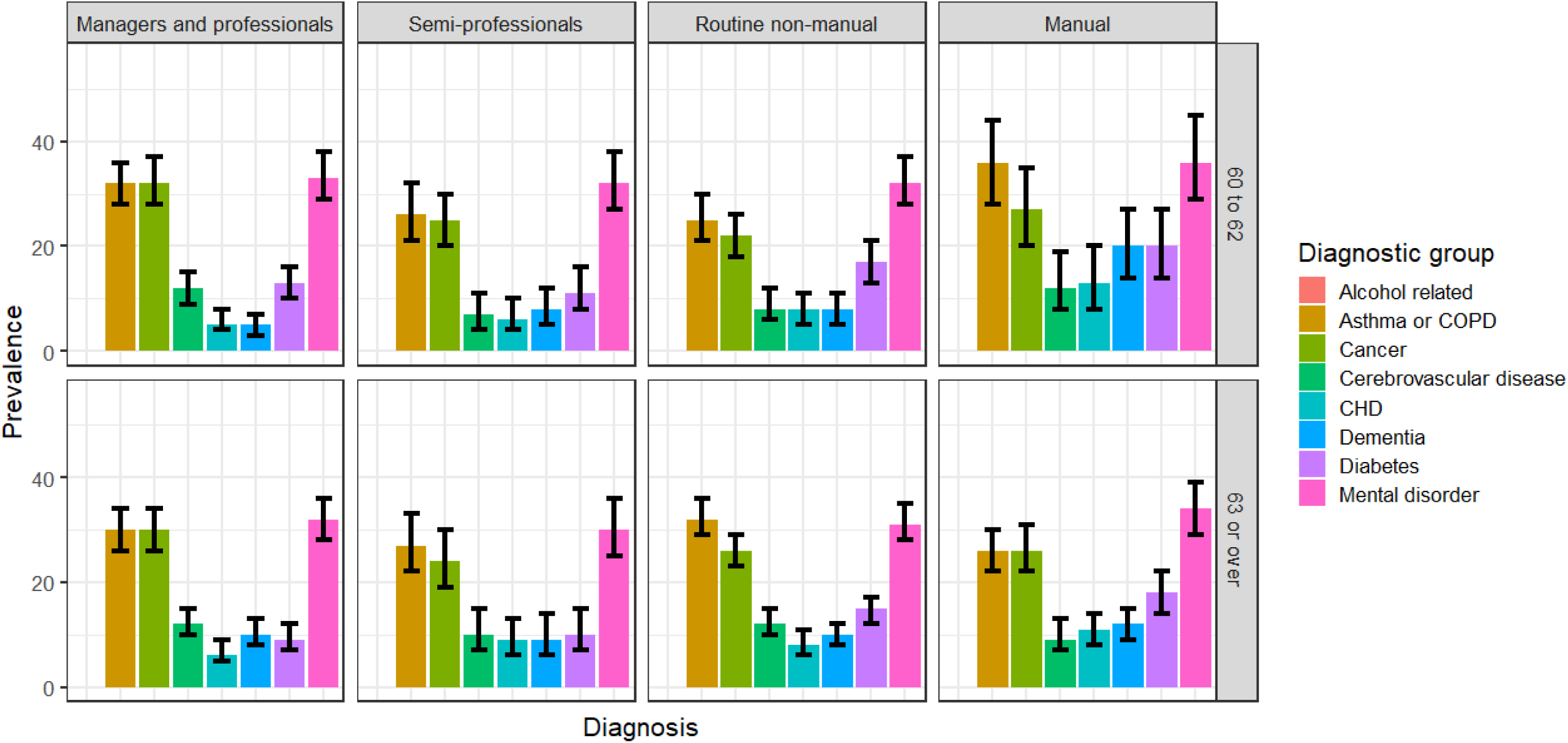
Prevalence rate of each diagnostic group of the major chronic illnesses on statutory retirement by occupational class and age at retirement, women (n=3291, average follow-up time 15.4 years). Results for diagnostic groups for which frequencies below five in the subgroup defined by occupational class and age at retirement could be deduced from the prevalence rate are omitted. Exact numbers can be seen in Appendix 2.

Among men, the most common diagnostic groups in most subgroups were mental disorders and cancers (Figure 3). No clear differences could be seen in the prevalence rates between the occupational classes or between those who retired earlier or later.

**Figure 3:**
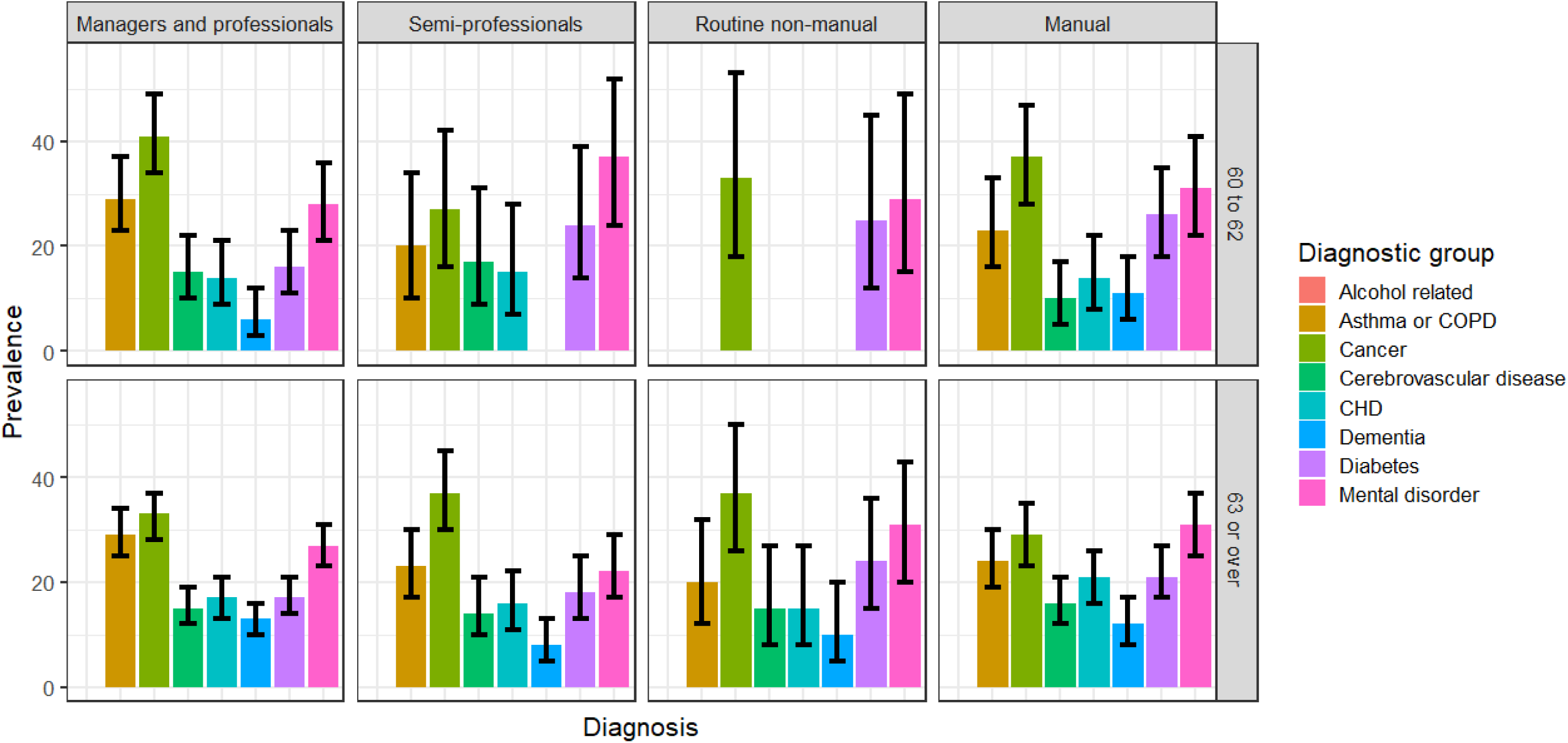
Prevalence rate of each diagnostic group of the major chronic illnesses on statutory retirement by occupational class and age at retirement, men (n=1268, average follow-up time 15.9 years). Results for diagnostic groups for which frequencies below five in the subgroup defined by occupational class and age at retirement could be deduced from the prevalence rate are omitted. Exact numbers can be seen in Appendix 2

## Discussion

Our results show clear differences in the number of healthy years on retirement by sex and age of retirement. Women have more healthy years on retirement than men, and those who retire early spend more healthy years on retirement than those who retire later. No clear occupational class gradient was observed in the number of healthy years spent on retirement. The difference between those with most healthy years on retirement and those with the least healthy years on retirement was 5.1 years. There were mostly small differences in the illnesses that occurred on retirement by occupational class, sex, or timing of retirement. However, diabetes was slightly more common among women from routine non-manual and manual classes regardless of retirement age and dementia among manual working women having retired early.

Most previous studies have included in their follow-up healthy years among individuals who are still working-aged. Therefore, direct comparisons to previous similar studies cannot be made, but we can nevertheless compare our results to previous studies on healthy ageing and retirement timing.

Previous studies on non-retired populations have consistently found higher socioeconomic position to be associated with healthy ageing (Wagg et al., 2021), as well as with having a higher expectancy of years lived without chronic diseases (Head et al., 2019). In contrast to previous studies, our study did not find consistent occupational class differences in healthy years on retirement. As occupational class is a socioeconomic indicator connected particularly to working life, it is possible that its effects diminish as the population enters retirement and as more years pass since occupational exposures.

However, this is unlikely to fully explain the discrepancy in the results, as the effects of occupational class do not disappear on retirement. In addition, our data are selective and consisted only of statutory retirees, thus those retired for disability are not included and consist of more people from lower occupational classes. As disability retirement is more common among those with detrimental health behaviours and work-related risk factors (Shiri et al., 2021), our sample of statutory retirees represents a comparatively healthy cohort of retirees to begin with.

The age of entering retirement decreased from the 1950s towards the end of the last century in the USA and Europe but has since then started to increase (Fisher et al., 2016). Most of the OECD countries have aimed to increase retirement ages through policy measures (*Pensions at a Glance 2023*, 2023). It is therefore likely that retirement ages continue to rise in most developed countries. While policies aiming to raise the retirement age may be justified from fiscal or labour market points of view, questions remain on what the impact of rising retirement age on the individuals and groups is, and how equal are the impacts on the individuals and groups. Our results indicate that those who retire later also enjoy less healthy years on retirement, as do men regardless of retirement age. Whether such unintended and unequitable consequences are an acceptable price to pay for the desired fiscal and labour force effects requires open discussion.

Certain factors render credibility to our results. Our data consist of a large number of participants identified reliably and comprehensively with no dropout from their former employer’s register. The health outcomes are derived from complete administrative registers and are all medically confirmed. However, not all possible medical diagnoses were included and the practical importance of a specific diagnosis to the individual was not known. For example, the cancer diagnoses range from local skin cancers to invasive non-curable conditions. The Royston-Parmar model, used to estimate healthy life-years in the combinations of sex, occupational class and age at retirement, estimates directly the baseline hazard function, and is therefore particularly suited for concrete predictions of healthy life years on retirement in absolute terms.

However, being based only on registers, our data permit only a limited number of contributing factors to be considered. For example, the contributions of health behaviours, further sociodemographic factors and working conditions to the examined healthy years on retirement could not be studied. The included socioeconomic variables may, however, partly act as proxies for health behaviours and working conditions (Lahelma et al., 2010)(Schram et al., 2021), and including them simultaneously could cause problems due to the strong overlap of the contributing factors.

In conclusion, our results indicate that there are clear disparities in the length of time spent without diseases during retirement. Policies aiming to raise retirement age should consider these disparities and weigh the desirable effects of increasing retirement age with possible negative and unequal effects on the individual.

## Data availability

The Helsinki Health Study survey data cannot be shared due to strict data protection laws and regulations. More information is available at the project website (helsinki.fi/hhs), in our data protection statement (helsinki.fi/hhs/data-protection-statement), and from. Permission to the secondary use of health and social data is granted by the Finnish Social and Health Data Permit Authority, Findata.

---

**Appendix 1:** Association of sex, occupational class, and age at retirement with predicted median years lived on statutory retirement without major chronic diseases, and predicted median age at first occurrence of illness.

| **Retirement age** | **Sex** | **SEP** | **n** | **Healthy years (95% CI)** | **Age at first illness (95% CI)** |
| --- | --- | --- | --- | --- | --- |
| 63 or over | Woman | Managers and Professionals | 543 | 9.0 (7.9–9.9) | 71.5 (69.3–73.1) |
| 63 or over | Woman | Semi-professionals | 243 | 9.8 (8.3–11.3) | 73 (71.2–74.8) |
| 63 or over | Woman | Routine non-manual | 666 | 8.5 (7.7–9.4) | 71.4 (70.3–72.5) |
| 63 or over | Woman | Manual | 388 | 9.1 (8.0–10.3) | 72.2 (70.9–73.6) |
| 63 or over | Man | Managers and Professionals | 389 | 8.2 (7.2–9.2) | 70.8 (69.9–72.0) |
| 63 or over | Man | Semi-professionals | 166 | 7.7 (6.3–9.2) | 70.8 (69.1–72.4) |
| 63 or over | Man | Routine non-manual | 59 | 6.5 (4.4–9.5) | 69.2 (63.0–71.4) |
| 63 or over | Man | Manual | 224 | 7.4 (6.0–8.7) | 69.8 (68.2–71.5) |
| 60-62 | Woman | Managers and Professionals | 470 | 9.7 (8.6–10.9) | 69.6 (68.2–70.8) |
| 60-62 | Woman | Semi-professionals | 251 | 9.4 (7.8–11.1) | 69.3 (67.2–71.8) |
| 60-62 | Woman | Routine non-manual | 393 | 11.6 (10.4–12.7) | 71.3 (69.6–72.9) |
| 60-62 | Woman | Manual | 137 | 9.3 (7.1–11.2) | 68.9 (65.8–71.5) |
| 60-62 | Man | Managers and Professionals | 143 | 9.4 (7.6–11.1) | 69.9 (68.1–71.4) |
| 60-62 | Man | Semi-professionals | 41 | 9.6 (5.9–13.9) | 68.1 (61.9–73.4) |
| 60-62 | Man | Routine non-manual | 24 | 7.5 (4.0–15.5) | 67.4 (60.8–75.0) |
| 60-62 | Man | Manual | 94 | 7.5 (5.3–9.6) | 66.4 (64.4–70.2) |

**Appendix 2:** Prevalence rate and 95% confidence interval of each diagnostic group of the first illness to end healthy years on retirement by occupational class and age at retirement by sex, SEP and retirement age. Results for diagnostic groups for which frequencies below five in the subgroup could be deduced from the prevalence rate are omitted.

| **Sex** | **SEP** | **Retirement age** | **Diagnostic group** | **Prevalence rate (95% CI)** |
| --- | --- | --- | --- | --- |
| Men | Managers and professionals | 63 or over | Diabetes | 17 (14–21) |
| Men | Managers and professionals | 60 to 62 | Diabetes | 16 (11–23) |
| Men | Semi-professionals | 63 or over | Diabetes | 18 (13–25) |
| Men | Semi-professionals | 60 to 62 | Diabetes | 24 (14–39) |
| Men | Routine non-manual | 63 or over | Diabetes | 24 (15–36) |
| Men | Routine non-manual | 60 to 62 | Diabetes | 25 (12–45) |
| Men | Manual | 63 or over | Diabetes | 21 (17–27) |
| Men | Manual | 60 to 62 | Diabetes | 26 (18–35) |
| Women | Managers and professionals | 63 or over | Diabetes | 9 (7–12) |
| Women | Managers and professionals | 60 to 62 | Diabetes | 13 (10–16) |
| Women | Semi-professionals | 63 or over | Diabetes | 10 (7–15) |
| Women | Semi-professionals | 60 to 62 | Diabetes | 11 (8–16) |
| Women | Routine non-manual | 63 or over | Diabetes | 15 (12–17) |
| Women | Routine non-manual | 60 to 62 | Diabetes | 17 (13–21) |
| Women | Manual | 63 or over | Diabetes | 18 (14–22) |
| Women | Manual | 60 to 62 | Diabetes | 20 (14–27) |
| Men | Managers and professionals | 63 or over | Asthma or COPD | 29 (25–34) |
| Men | Managers and professionals | 60 to 62 | Asthma or COPD | 29 (23–37) |
| Men | Semi-professionals | 63 or over | Asthma or COPD | 23 (17–30) |
| Men | Semi-professionals | 60 to 62 | Asthma or COPD | 20 (10–34) |
| Men | Routine non-manual | 63 or over | Asthma or COPD | 20 (12–32) |
| Men | Routine non-manual | 60 to 62 | Asthma or COPD | Omitted |
| Men | Manual | 63 or over | Asthma or COPD | 24 (19–30) |
| Men | Manual | 60 to 62 | Asthma or COPD | 23 (16–33) |
| Women | Managers and professionals | 63 or over | Asthma or COPD | 30 (26–34) |
| Women | Managers and professionals | 60 to 62 | Asthma or COPD | 32 (28–36) |
| Women | Semi-professionals | 63 or over | Asthma or COPD | 27 (22–33) |
| Women | Semi-professionals | 60 to 62 | Asthma or COPD | 26 (21–32) |
| Women | Routine non-manual | 63 or over | Asthma or COPD | 32 (29–36) |
| Women | Routine non-manual | 60 to 62 | Asthma or COPD | 25 (21–30) |
| Women | Manual | 63 or over | Asthma or COPD | 26 (22–30) |
| Women | Manual | 60 to 62 | Asthma or COPD | 36 (28–44) |
| Men | Managers and professionals | 63 or over | Dementia | 13 (10–16) |
| Men | Managers and professionals | 60 to 62 | Dementia | 6 (3–12) |
| Men | Semi-professionals | 63 or over | Dementia | 8 (5–13) |
| Men | Semi-professionals | 60 to 62 | Dementia | Omitted |
| Men | Routine non-manual | 63 or over | Dementia | 10 (5–20) |
| Men | Routine non-manual | 60 to 62 | Dementia | Omitted |
| Men | Manual | 63 or over | Dementia | 12 (8–17) |
| Men | Manual | 60 to 62 | Dementia | 11 (6–18) |
| Women | Managers and professionals | 63 or over | Dementia | 10 (8–13) |
| Women | Managers and professionals | 60 to 62 | Dementia | 5 (3–7) |
| Women | Semi-professionals | 63 or over | Dementia | 9 (6–14) |
| Women | Semi-professionals | 60 to 62 | Dementia | 8 (5–12) |
| Women | Routine non-manual | 63 or over | Dementia | 10 (8–12) |
| Women | Routine non-manual | 60 to 62 | Dementia | 8 (5–11) |
| Women | Manual | 63 or over | Dementia | 12 (9–15) |
| Women | Manual | 60 to 62 | Dementia | 20 (14–27) |
| Men | Managers and professionals | 63 or over | Mental disorder | 27 (23–31) |
| Men | Managers and professionals | 60 to 62 | Mental disorder | 28 (21–36) |
| Men | Semi-professionals | 63 or over | Mental disorder | 22 (17–29) |
| Men | Semi-professionals | 60 to 62 | Mental disorder | 37 (24–52) |
| Men | Routine non-manual | 63 or over | Mental disorder | 31 (20–43) |
| Men | Routine non-manual | 60 to 62 | Mental disorder | 29 (15–49) |
| Men | Manual | 63 or over | Mental disorder | 31 (25–37) |
| Men | Manual | 60 to 62 | Mental disorder | 31 (22–41) |
| Women | Managers and professionals | 63 or over | Mental disorder | 32 (28–36) |
| Women | Managers and professionals | 60 to 62 | Mental disorder | 33 (29–38) |
| Women | Semi-professionals | 63 or over | Mental disorder | 30 (25–36) |
| Women | Semi-professionals | 60 to 62 | Mental disorder | 32 (27–38) |
| Women | Routine non-manual | 63 or over | Mental disorder | 31 (28–35) |
| Women | Routine non-manual | 60 to 62 | Mental disorder | 32 (28–37) |
| Women | Manual | 63 or over | Mental disorder | 34 (29–39) |
| Women | Manual | 60 to 62 | Mental disorder | 36 (29–45) |
| Men | Managers and professionals | 63 or over | CHD | 17 (13–21) |
| Men | Managers and professionals | 60 to 62 | CHD | 14 (9–21) |
| Men | Semi-professionals | 63 or over | CHD | 16 (11–22) |
| Men | Semi-professionals | 60 to 62 | CHD | 15 (7–28) |
| Men | Routine non-manual | 63 or over | CHD | 15 (8–27) |
| Men | Routine non-manual | 60 to 62 | CHD | Omitted |
| Men | Manual | 63 or over | CHD | 21 (16–26) |
| Men | Manual | 60 to 62 | CHD | 14 (8–22) |
| Women | Managers and professionals | 63 or over | CHD | 6 (5–9) |
| Women | Managers and professionals | 60 to 62 | CHD | 5 (4–8) |
| Women | Semi-professionals | 63 or over | CHD | 9 (6–13) |
| Women | Semi-professionals | 60 to 62 | CHD | 6 (4–10) |
| Women | Routine non-manual | 63 or over | CHD | 8 (6–11) |
| Women | Routine non-manual | 60 to 62 | CHD | 8 (5–11) |
| Women | Manual | 63 or over | CHD | 11 (8–14) |
| Women | Manual | 60 to 62 | CHD | 13 (8–20) |
| Men | Managers and professionals | 63 or over | Cerebrovascular disease | 15 (12–19) |
| Men | Managers and professionals | 60 to 62 | Cerebrovascular disease | 15 (10–22) |
| Men | Semi-professionals | 63 or over | Cerebrovascular disease | 14 (10–21) |
| Men | Semi-professionals | 60 to 62 | Cerebrovascular disease | 17 (9–31) |
| Men | Routine non-manual | 63 or over | Cerebrovascular disease | 15 (8–27) |
| Men | Routine non-manual | 60 to 62 | Cerebrovascular disease | Omitted |
| Men | Manual | 63 or over | Cerebrovascular disease | 16 (12–21) |
| Men | Manual | 60 to 62 | Cerebrovascular disease | 10 (5–17) |
| Women | Managers and professionals | 63 or over | Cerebrovascular disease | 12 (10–15) |
| Women | Managers and professionals | 60 to 62 | Cerebrovascular disease | 12 (9–15) |
| Women | Semi-professionals | 63 or over | Cerebrovascular disease | 10 (7–15) |

| **Sex** | **SEP** | **Retirement**<br>**age** | **Diagnostic group** | **Prevalence rate**<br>**(95% CI)** |
| --- | --- | --- | --- | --- |
| Women | Semi-professionals | 60 to 62 | Cerebrovascular disease | 7 (4–11) |
| Women | Routine non-manual | 63 or over | Cerebrovascular disease | 12 (10–15) |
| Women | Routine non-manual | 60 to 62 | Cerebrovascular disease | 8 (6–12) |
| Women | Manual | 63 or over | Cerebrovascular disease | 9 (7–13) |
| Women | Manual | 60 to 62 | Cerebrovascular disease | 12 (8–19) |
| Men | Managers and professionals | 63 or over | Cancer | 33 (28–37) |
| Men | Managers and professionals | 60 to 62 | Cancer | 41 (34–49) |
| Men | Semi-professionals | 63 or over | Cancer | 37 (30–45) |
| Men | Semi-professionals | 60 to 62 | Cancer | 27 (16–42) |
| Men | Routine non-manual | 63 or over | Cancer | 37 (26–50) |
| Men | Routine non-manual | 60 to 62 | Cancer | 33 (18–53) |
| Men | Manual | 63 or over | Cancer | 29 (23–35) |
| Men | Manual | 60 to 62 | Cancer | 37 (28–47) |
| Women | Managers and professionals | 63 or over | Cancer | 30 (26–34) |
| Women | Managers and professionals | 60 to 62 | Cancer | 32 (28–37) |
| Women | Semi-professionals | 63 or over | Cancer | 24 (19–30) |
| Women | Semi-professionals | 60 to 62 | Cancer | 25 (20–30) |
| Women | Routine non-manual | 63 or over | Cancer | 26 (23–29) |
| Women | Routine non-manual | 60 to 62 | Cancer | 22 (18–26) |
| Women | Manual | 63 or over | Cancer | 26 (22–31) |
| Women | Manual | 60 to 62 | Cancer | 27 (20–35) |
| Men | Managers and professionals | 63 or over | Alcohol related | Omitted |
| Men | Managers and professionals | 60 to 62 | Alcohol related | Omitted |
| Men | Semi-professionals | 63 or over | Alcohol related | Omitted |
| Men | Semi-professionals | 60 to 62 | Alcohol related | Omitted |
| Men | Routine non-manual | 63 or over | Alcohol related | Omitted |
| Men | Routine non-manual | 60 to 62 | Alcohol related | Omitted |
| Men | Manual | 63 or over | Alcohol related | Omitted |
| Men | Manual | 60 to 62 | Alcohol related | Omitted |
| Women | Managers and professionals | 63 or over | Alcohol related | Omitted |
| Women | Managers and professionals | 60 to 62 | Alcohol related | Omitted |
| Women | Semi-professionals | 63 or over | Alcohol related | Omitted |
| Women | Semi-professionals | 60 to 62 | Alcohol related | Omitted |
| Women | Routine non-manual | 63 or over | Alcohol related | Omitted |
| Women | Routine non-manual | 60 to 62 | Alcohol related | Omitted |
| Women | Manual | 63 or over | Alcohol related | Omitted |
| Women | Manual | 60 to 62 | Alcohol related | Omitted |

## References

Dalstra, J., Kunst, A., Borrell, C., Breeze, E., Cambois, E., Costa, G., Geurts, J., Lahelma, E., Van Oyen, H., Rasmussen, N., Regidor, E., Spadea, T., & Mackenbach, J. (2005). Socioeconomic differences in the prevalence of common chronic diseases: An overview of eight European countries. International Journal of Epidemiology, 34(2), 316–326. 10.1093/ije/dyh386

Fisher, G. G., Chaffee, D. S., & Sonnega, A. (2016). Retirement Timing: A Review and Recommendations for Future Research. Work, Aging and Retirement, 2(2), 230–261. 10.1093/workar/waw001

Garrouste, C., & Perdrix, E. (2022). Is there a consensus on the health consequences of retirement? A literature review. Journal of Economic Surveys, 36(4), 841–879. 10.1111/joes.12466

GBD 2015 DALYs and HALE Collaborators. (2016). Global, Regional, and National Disability-adjusted Life Years (Dalys) for 315 Diseases and Injuries and Healthy Life Expectancy (Hale), 1990-2015: A Systematic Analysis for the Global Burden of Disease Study 2015. https://hdl.handle.net/10292/10072

Global Burden of Disease 2019 Cancer Collaboration. (2022). Cancer Incidence, Mortality, Years of Life Lost, Years Lived With Disability, and Disability-Adjusted Life Years for 29 Cancer Groups From 2010 to 2019: A Systematic Analysis for the Global Burden of Disease Study 2019. JAMA Oncology, 8(3), 420–444. 10.1001/jamaoncol.2021.6987

Haukka, J. (2004). Finnish health and social welfare registers in epidemiological research. Norsk Epidemiologi, 14(1), Article 1. 10.5324/nje.v14i1.284

Head, J., Chungkham, H. S., Hyde, M., Zaninotto, P., Alexanderson, K., Stenholm, S., Salo, P., Kivimäki, M., Goldberg, M., Zins, M., Vahtera, J., & Westerlund, H. (2019). Socioeconomic differences in healthy and disease-free life expectancy between ages 50 and 75: A multi-cohort study. European Journal of Public Health, 29(2), 267–272. 10.1093/eurpub/cky215

Heald, A. H., Stedman, M., Davies, M., Livingston, M., Alshames, R., Lunt, M., Rayman, G., & Gadsby, R. (2020). Estimating life years lost to diabetes: Outcomes from analysis of National Diabetes Audit and Office of National Statistics data. Cardiovascular Endocrinology & Metabolism, 9(4), 183. 10.1097/XCE.0000000000000210

Jackson, C., Metcalfe, P., Amdahl, J., Warkentin, M. T., Sweeting, M., & Kunzmann, K. (2024). flexsurv: Flexible Parametric Survival and Multi-State Models (Version 2.3.2) [Computer software]. https://cran.r-project.org/web/packages/flexsurv/index.html

Junna, L., Tarkiainen, L., Leinonen, T., Korhonen, K., & Martikainen, P. (2024, October 23). Trends in working life expectancy by education and occupational social class in Finland, 1991–2020. Finnish Centre for Pensions. https://www.julkari.fi/handle/10024/149888

Lahelma, E., Lallukka, T., Laaksonen, M., Martikainen, P., Rahkonen, O., Chandola, T., Head, J., Marmot, M., Kagamimori, S., Tatsuse, T., & Sekine, M. (2010). Social class differences in health behaviours among employees from Britain, Finland and Japan: The influence of psychosocial factors. Health & Place, 16(1), 61–70. 10.1016/j.healthplace.2009.08.004

Ma, H., Wang, X., Xue, Q., Li, X., Liang, Z., Heianza, Y., Franco, O. H., & Qi, L. (2023). Cardiovascular Health and Life Expectancy Among Adults in the United States. Circulation, 147(15), 1137–1146. 10.1161/CIRCULATIONAHA.122.062457

Ng, R., Kornas, K., Sutradhar, R., Wodchis, W. P., & Rosella, L. C. (2018). The current application of the Royston-Parmar model for prognostic modeling in health research: A scoping review. Diagnostic and Prognostic Research, 2(1), 4. 10.1186/s41512-018-0026-5

Nyberg, S. T., Batty, G. D., Pentti, J., Madsen, I. E. H., Alfredsson, L., Bjorner, J. B., Borritz, M., Burr, H., Ervasti, J., Goldberg, M., Jokela, M., Knutsson, A., Koskinen, A., Lallukka, T., Lindbohm, J. V., Nielsen, M. L., Oksanen, T., Pejtersen, J. H., Pietiläinen, O.,…Kivimäki, M. (2022). Association of alcohol use with years lived without major chronic diseases: A multicohort study from the IPD-Work consortium and UK Biobank. The Lancet Regional Health - Europe, 19, 100417. 10.1016/j.lanepe.2022.100417

Nyberg, S. T., Singh-Manoux, A., Pentti, J., Madsen, I. E. H., Sabia, S., Alfredsson, L., Bjorner, J. B., Borritz, M., Burr, H., Goldberg, M., Heikkilä, K., Jokela, M., Knutsson, A., Lallukka, T., Lindbohm, J. V., Nielsen, M. L., Nordin, M., Oksanen, T., Pejtersen, J. H.,…Kivimäki, M. (2020). Association of Healthy Lifestyle With Years Lived Without Major Chronic Diseases. JAMA Internal Medicine, 180(5), 760–768. 10.1001/jamainternmed.2020.0618

Pensions at a Glance 2023. (2023, December 13). OECD. https://www.oecd.org/en/publications/pensions-at-a-glance-2023_678055dd-en.html

R Core Team. (2023). R: A Language and Environment for Statistical Computing (Version 4.3.1) [Computer software]. R Foundation for Statistical Computing. https://www.R-project.org/

Royston, P., & Parmar, M. K. B. (2002). Flexible parametric proportional-hazards and proportional-odds models for censored survival data, with application to prognostic modelling and estimation of treatment effects. Statistics in Medicine, 21(15), 2175–2197. 10.1002/sim.1203

Schram, J. L., Solovieva, S., Leinonen, T., Viikari-Juntura, E., Burdorf, A., & Robroek, S. J. (2021). The influence of occupational class and physical workload on working life expectancy among older employees. Scandinavian Journal of Work, Environment & Health, 47(1), 5–14. 10.5271/sjweh.3919

Shiri, R., Hiilamo, A., & Lallukka, T. (2021). Indicators and determinants of the years of working life lost: A narrative review. Scandinavian Journal of Public Health, 49(6), 666–674. 10.1177/1403494821993669

Society at a Glance 2019. (2019, March 27). OECD. https://www.oecd.org/en/publications/society-at-a-glance-2019_soc_glance-2019-en.html

Wagg, E., Blyth, F. M., Cumming, R. G., & Khalatbari-Soltani, S. (2021). Socioeconomic position and healthy ageing: A systematic review of cross-sectional and longitudinal studies. Ageing Research Reviews, 69, 101365. 10.1016/j.arr.2021.101365

